# Speaker Role Identification in Clinical Conversations^*^

**DOI:** 10.1101/2025.08.14.25332837

**Authors:** Andrew Zolensky, Kuk Jin Jang, Janice Sabin, Andrea Hartzler, Basam Alasaly, Sriharsha Mopidevi, Mark Liberman, Kevin Johnson

## Abstract

Patient-clinician communication research is crucial for understanding interaction dynamics and for predicting outcomes that are associated with clinical discourse. Traditionally, interaction analysis is conducted manually because of challenges such as Speaker Role Identification (SRI), which must reliably differentiate between doctors, medical assistants, patients, and other caregivers in the same room. Although automatic speech recognition with diarization can efficiently create a transcript with separate labels for each speaker, these systems are not able to assign roles to each person in the interaction. Previous SRI studies in task-oriented scenarios have directly predicted roles using linguistic features, bypassing diarization. However, to our knowledge nobody has investigated SRI in clinical settings. We explored whether Large Language Models (LLMs) such as BERT could accurately identify speaker roles in clinical transcripts, with and without diarization. We used veridical turn segmentation and diarization identifiers, fine-tuning each model at varying levels of identifier corruption to assess impact on performance. Our results demonstrate that BERT achieves high performance with linguistic signals alone (82% accuracy/82% F1-score), while incorporating accurate diarization identifiers further enhances accuracy (95%/95%). We conclude that fine-tuned LLMs are effective tools for SRI in clinical settings.

## 1. Introduction

Patient-clinician interaction forms a central component of healthcare, generating medically relevant data and situating interventions. Effective communication between clinicians and patients impacts healthcare utilization, medical adherence, and various other positive and negative functional outcomes [1][2][3]. Health systems recognize the critical role of communication by adopting research-informed guidelines for judging quality of patient-clinician communication, such as the Six Core Competencies for medical professionals defined by the Accreditation Council for Graduate Medical Education [4][5]. One such competency is Interpersonal and Communication Skills (ICS), including “effective information exchange and teaming with patients, their families, and other health professionals.” Similarly, effective communication is central to Patient-Centered Care (PCC), a model in which patient participation and decision-sharing are key [3][6]. Nevertheless, precisely evaluating what constitutes “effective communication” is an ongoing research challenge [7].

Evaluating communication requires electronic representations of recorded conversations, which can be provided as textual transcripts derived from speech-to-text applications. For these transcripts to be useful for further research, it is necessary to accurately attribute each utterance to the correct participant, a process we refer to as Speaker Role Identification (SRI). Typical roles in a clinical setting include “physician,” “medical assistant,” and “patient.”

Diarization tools, often included with automated transcription services, label audio by distinguishing between speakers based on acoustic features. To do so, diarization pipelines segment audio into turns using voice activity detection and then utilize clustering algorithms to group utterances based on spectrographic representations [8]. However, these tools assign generic diarization identifiers such as “Speaker 1” or “Speaker 2,” without mapping them to functional roles like “physician” or “patient”. As a result, SRI researchers have employed pipelines that use diarization identifiers to group utterances into disjoint sets, then employ either structural heuristics or lexical modeling on each group to infer speaker roles [9][10]. However, diarization errors may propagate through these pipelines and significantly undermine the accuracy of SRI.

Furthermore, reliance on audio cues causes diarization algorithms to overlook valuable linguistic information present in the transcript, which might be useful for downstream SRI, such as role-specific vocabulary. Research on SRI in parallel sources of conversational data, such as call-center dialogues [9], Air Traffic Control [11], and at-home care [12], suggests that roles may be robustly identified solely from the textual content of transcribed utterances using Support Vector Machines [12], Bi-LSTMs, CNNs, and Transformers [9][11], without the use of any diarization identifiers. Transformers can attend to previous utterance context in addition to the target utterance, allowing for more flexible utterance grouping strategies. However, these studies overlook the possibility that diarization identifiers can be useful as categorical inputs to models.

Our primary aim is to test the viability of SRI in the context of clinical conversations by using Large Language Models (LLMs), as these have shown improvements in a broad set of natural language processing (NLP) tasks over traditional, task-specific architectures [18], and have been used successfully for SRI in other domains [19]. We fine-tune BERT, an LLM based on the encoder-only transformer architecture [18], to perform SRI on a conversational dataset of over 25,000 utterances from doctors, patients, medical assistants, and others, made during recorded, out-patient clinical visits. To establish baselines for comparison, we also train decision trees and gradient boosted decision trees. Additionally, we assess the effect of varying multiple design hyperparameters that have been evaluated in the SRI literature, including use of heuristic and linguistic features, and assess the impact of various contextual grouping strategies based on proximity to target utterance and use of diarization identifiers. Lastly, we quantify the performance impact of diarization use and error for each method.

## 2. Previous Works

SRI has been applied to several domains over the years but has seen limited references in clinical informatics. Most SRI applications relate to social signal processing and arise in studies of task-specific communication. In each of these domains, SRI has been motivated by the need for richer representation of conversational datasets. It generally supports indexing and retrieval functions for large human conversation datasets [16].

Early work on SRI was done on conversational data for radio broadcast, television news, and talk shows [13][14][15]. Other SRI work has occurred in domains such as Air Traffic Control, where it has been used to differentiate between pilots and ATC officials [11]. Recently, SRI has also been applied to differentiate between social roles in work-related meetings [16], to distinguish between assistant and customer in call-center conversations [9], and to distinguish between study participants and administrators in conversations related to clinical trials for autism spectrum disorder [10]. Lastly, in at-home health settings, SRI has been applied successfully to predict caregiver and patient roles [12].

SRI has been approached using several feature representations. The best-performing models to date utilize features extracted via Automated Speech Recognition (ASR) software and other structural features from conversations [12][14][15], spectrographic and prosodic features [11][16], and in some cases video signals as well [13]. However, it remains possible to achieve high performance using text alone [9][11][12]. SRI has also been performed using a variety of methods mentioned previously.

Research exists in non-clinical domains, including at-home care settings, but SRI has not yet been applied in clinical conversations between doctors and patients. Additionally, the possibility of incorporating diarization identifiers remains underexplored. Most models ignore diarization identifiers, or use them for grouping utterances, without allowing downstream models to directly attend to them.

From an implementation standpoint, the text-only transformer model is the simplest approach that preserves the ability to fit arbitrarily complex semantic patterns. We hypothesize that lexical signals alone are sufficient to identify speaker roles in clinical visit conversations, and that inclusion of diarization identifiers as model inputs will further improve performance. To test this, we applied BERT to a clinical conversation dataset, evaluating its effectiveness in identifying the roles of patient, doctor, and medical assistant.

## 3. Materials & Methods

### 3.1 Task Definition

The task of SRI consists of accurately identifying one speaker role label for each utterance in a recorded conversation. We define a conversation as a textual transcript consisting of a list of utterances. Each utterance is represented as a sequence of tokens, and every token is drawn from a specified vocabulary. Each utterance corresponds to one a speaker role, which in our case is one of: “patient,” “doctor,” “medical assistant,” or “other.” The task of SRI is, given a textual transcript, uncover the speaker role associated with each utterance.

Although it is possible to use the entire conversation as context for each utterance, in this study, we consider only previous utterances within a specified number of utterances of the target utterance as context. A sliding window, which we call a “context window”, is applied to ease computation and ensure inputs fit fully within LLM context limits. Restriction to the previous context is done to leave it unambiguous which is the target utterance (the last utterance in the window). The number of utterances in the window, including the target utterance, is called the “context size”. This is treated as an independent variable in our experiments.

### 3.2 Dataset

We used 117 manually transcribed clinical visit transcripts from the Establishing Focus (EF) Study [17], comprising a total of 27,505 usable utterances across 117 patients and 27 providers, each labeled with speaker role (doctor, patient, medical assistant, and others). The EF study was a randomized control trial designed to assess the impact of specialized clinician training in agenda-setting on communication behaviors and functional outcomes of clinical visits. Participating clinicians were trained and had their visits audio-taped six months afterwards (March 2004-March 2005). Recordings spanned 1460 patients and 48 clinicians. The clinical population was generated via convenience sampling of 12 community-based primary care clinics in the Puget Sound area of Washington. Consented patients were over the age of 18 and had previous visits with the clinician. Encounters shorter than three minutes were removed from the dataset. Encounters lasted an average duration of 32 minutes, 58 seconds (std. dev. = 14 minutes, 40 seconds).

### 3.3 Pre-Processing

Transcript pre-processing was performed using the Python docx module, version 1.2.0. The steps are shown in Figure 1. An example of a transcript prior to processing is shown in Figure 2. Human transcribers inserted paragraph breaks between successive transcribed speaker turns, which consist of a speaker label and an utterance, allowing them to be easily extracted using the docx module. For each extracted turn, we normalized spacing by replacing tab characters with a single white space and stripping leading and trailing whitespaces from the result. Next, if a turn began with an open parentheses or bracket, we dropped it from the transcript, as it was most likely a description of non-verbal activity, such as “(MA enters the room)”. To separate the utterance and the speaker label, we split each remaining turn on the colon character (“:”). If this failed to extract two separate items for a given utterance, we attempted to split on semi-colons (“;”), and lastly on spaces (“”), in case transcribers forgot to include relevant punctuation. If all these methods failed, the turn was dropped from the transcript. If speaker label extraction succeeded, we removed all non-alphanumeric characters from the speaker label, stripped trailing and leading whitespaces, and converted it to lowercase before saving both the speaker label and the utterance.

**Fig. 1.**
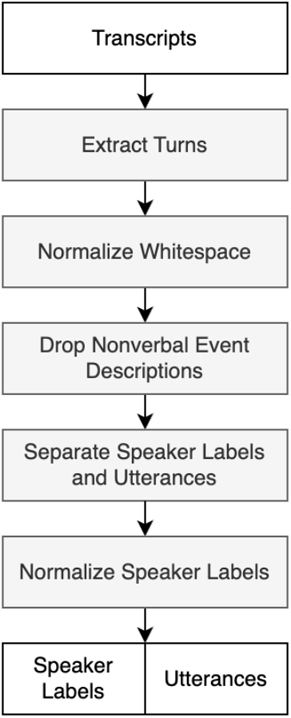
Diagram displaying pre-processing steps taken for transcript data prior to modeling.

**Fig. 2.**
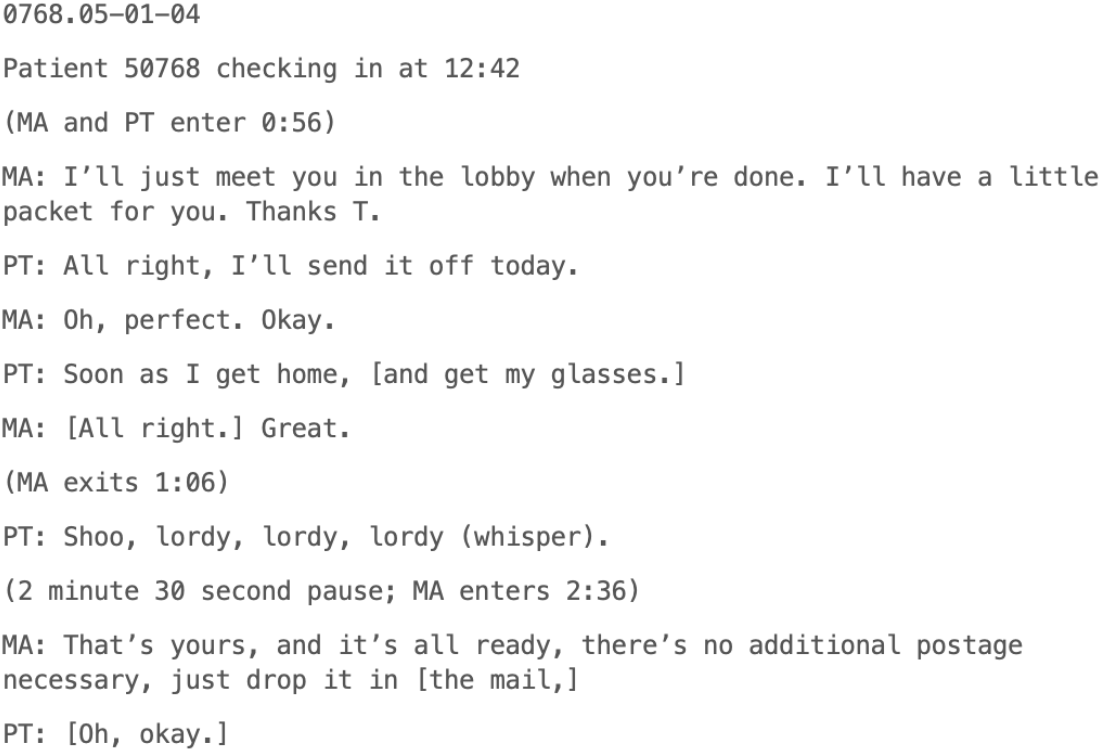
An example transcript from the EF dataset. Only the beginning is shown. Identifiable information has been replaced with dummy values.

Next, we collected all extracted speaker labels across our corpus, removed duplicates, and manually pruned the results to remove malformed labels (e.g. (e.g., “Case ID Number,” “Stop Time,” and “Patient”, the latter likely coming from lines such as “Patient 50768 checking in at 12:42,” rather than from actual utterance labels, which would have been coded as “PT”). For malformed labels, we also excluded the associated utterances from our dataset. We then categorized each distinct speaker label in the resulting set to one of “patient,” “doctor,” “medical assistant,” or “other”, and applied this mapping to our dataset to normalize the speaker labels.

To compute features for each utterance in each transcript, we computed the “line number,” or number of preceding utterances in the transcript plus one, for that utterance, and stored it along with the utterance content and the speaker role. Additionally, we computed the “second-person pronoun proportion,” the percentage of pronouns from that utterance that were second person, as opposed to first or third person. This was computed by searching for exact matches between a listed set of pronouns and the words in each utterance.

### 3.4 Diarization Error Simulation

We investigated whether numeric speaker identities computed via diarization software would provide models with helpful signals for reliably identifying speaker roles throughout transcripts, and if so, to what degree errors in diarization pipelines would adversely impact downstream SRI performance. Rather than relying on open source diarization tools that would provide us with a definite accuracy, we used our ground truth labels to simulate diarization at varying degrees of corruption. We did so by assigning each speaker role within a transcript to a numeric identifier and then randomly reassigning a pre-specified percentage of the numeric identifiers from the pool of identifiers in the conversation. We refer to this percentage as the “error rate”. The error rates we tested were 0%, 10%, 25%, and 50%.

### 3.5 Models

Three main model types were evaluated:

- Gradient-boosted decision tree classifiers using the Gini criterion as purity measure/objective function, a max depth of two, and 100 estimators, trained on the proportion of second-person pronouns in each utterance, as well as the utterance line number.
- Gradient-boosted decision tree classifiers using the Gini criterion as purity measure/objective function, a max depth of five, and 100 estimators, learning rate of 0.1, trained on the lexical content of the utterance using a bag of words approach.
- BERT-base-cased models with a multiclass classification head fine-tuned on the utterance content (sequence data) with utterance line number appended at the end, to classify speaker role.

We refer to instances of our first model type as “heuristics-based” models, and instances of the second and third types as “linguistic-based” models. For both the heuristics-based and linguistic-based boosted decision trees, we trained decision trees with identical max depth and purity settings for comparison.

All tree-based models were trained using scikit-learn, version 1.6.1. For BERT models, the tokenizer, model, and configuration files were loaded from the Hugging Face Transformers Library, version 4.48.0. The models were fine-tuned using the Hugging Face Trainer class.

All models were evaluated under differing context sizes, parametrized by the number of preceding utterances. Models received a sliding window of utterances and predicted the speaker of the final utterance. Inputs to BERT models were trimmed to avoid exceeding the token limit of 512 tokens for the model.

### 3.6 Overview of Experiments

Our primary aim was to evaluate the effectiveness of LLMs for SRI in clinical conversations. We also compared several methodological choices from the SRI literature, such heuristics-based versus linguistic-based models, basic machine learning versus LLMs, use of diarization identifiers, and using diarization identifiers to group utterances versus as direct model inputs.

To compare heuristic and linguistic approaches, we trained decision trees and boosted decision trees under two schemes: (1) models using bag-of-words features, and (2) models using simple heuristics; namely, pronoun proportion and utterance line number.

To evaluate the use of diarization identifiers for BERT, we experimented with three separate approaches, each attending to only a “context window” of previous utterances:

- BERT with no additional speaker information.
- BERT where the context window is filtered to only include utterances with the same diarization identifier as the target utterance. We call this a “grouping-based” model.
- BERT where the diarization identifier for each utterance is prefixed to that utterance, in the form “speaker<identifier>: “. We call this a “token-based” model.

This allowed us to compare LLM models without diarization, LLMs with grouped context based on speaker, and LLMs with diarization identifiers as tokens.

For tree-based models, diarization was handled differently, as the token-based strategy does not apply to bag-of-words features. Tree models which did not use diarization only attended to the target utterance without conversational context. For all other tree models, a grouping-based strategy was used. For heuristics-based trees, feature values were averaged over utterances from the same speaker within the context window; for linguistic-based trees, features were aggregated as the union of all words used by the same speaker within the context window.

### 3.7 Training and Optimization

Our dataset was split into training (70%), validation (15%), and testing (15%) subsets at the level of conversation id prior to all experiments. Proportion of utterances due to patients, doctors, medical assistants, and others in the dataset was 49%, 40%, 11%, and 0% (0.001% unrounded), respectively, and was preserved in each subset. All models were trained for exactly one epoch. BERT models were trained using the AdamW optimizer with a learning rate of 5e-05, and batch size of 32. No prompts were used.

### 3.8 Evaluation

Performance was assessed using accuracy and weighted F1 score on the testing dataset averaged across all speaker categories. Accuracy was computed as the number of ground truth labels predicted correctly. The F1 score, computed as the harmonic mean of precision and recall for a single class, was averaged across classes using weights proportional to the support to obtain the weighted F1 score. The weighted F1 score was chosen to aggregate per-class performance in a way that reflects the real-world distribution of utterances by speaker role in clinical environments.

### 3.9 Computational Environment

All experiments were performed on a dedicated GPU compute cluster, equipped with 4 vCPUs, 28 GB system memory, and a single NVIDIA T4 GPU (16 GB GDDR6). The software stack included Databricks Runtime 17.0.

### 3.10 Ethics

This study utilized transcripts containing protected health information (PHI) from the EF dataset [17], under a Data Use Agreement between the University of Washington (UW) and the University of Pennsylvania (UPenn). The protocol was approved by the UW IRB (STUDY000005436), listing UPenn as the relying institution via the SMART Master Reliance Agreement. All data were stored in secure, access-controlled environments only accessible to authorized study personnel. All handling of PHI was conducted in compliance with HIPAA and relevant institutional policies, as reflected in the IRB determination (exempt). Re-consent of participants was not required per IRB approval. Data used and machine learning models developed in this work are not publicly released to maintain the safety of participant data.

## 4. Results

### 4.1 Model Class

Accuracy and F1 scores for all models are shown in Table 1. The BERT models scored higher than the decision trees and boosted trees with respect to both accuracy and F1 for each setting of context size and diarization error rate. BERT models achieved balanced accuracy and F1 scores. Tree models achieved mostly balanced scores, but many of them displayed F1 scores anywhere from 2-6% lower than corresponding accuracy scores.

**Table 1.** Accuracy and F1 scores for BERT, decision-tree, and gradient-boosted decision tree models across alternative input and experimental hyperparameter settings. The column “Diarization Use” indicates whether grouping-based or token-based methods were used. “Inputs” indicates whether heuristics-based features or linguistic-based features were used, the latter encompassing bag of words features and text sequences. “Context Size” indicates the number of preceding utterances, including the target utterance, that were used in the context window. “Diarization Error” refers to experiments that used diarization and varying error levels, and the “No Diarization” refers to models that do not use diarization.

| Model | Diarization Use | Inputs | Context Size | Diarization Error |  |  |  | No Diarization |
| --- | --- | --- | --- | --- | --- | --- | --- | --- |
|  |  |  |  | 50% error | 25% error | 10% error | 0% error |  |
| Decision Tree | N/A | Heuristics | 1 | N/A | N/A | N/A | N/A | 0.62/0.58 |
| Decision Tree | Grouped | Heuristics | 2 | 0.58/0.54 | 0.60/0.56 | 0.61/0.58 | 0.62/0.59 | N/A |
| Decision Tree | Grouped | Heuristics | 5 | 0.62/0.61 | 0.67/0.66 | 0.70/0.70 | 0.71/0.71 | N/A |
| Decision Tree | Grouped | Heuristics | 10 | 0.64/0.63 | 0.70/0.69 | 0.74/0.73 | **0.76/0.76** | N/A |
| Decision Tree | N/A | Bag of Words | 1 | N/A | N/A | N/A | N/A | 0.62/0.57 |
| Decision Tree | Grouped | Bag of Words | 2 | 0.55/0.51 | 0.58/0.54 | 0.60/0.55 | 0.62/0.56 | N/A |
| Decision Tree | Grouped | Bag of Words | 5 | 0.58/0.56 | 0.62/0.60 | 0.67/0.64 | 0.71/0.68 | N/A |
| Decision Tree | Grouped | Bag of Words | 10 | 0.55/0.53 | 0.62/0.59 | 0.68/0.66 | **0.73/0.71** | N/A |
| Boosted Trees | N/A | Heuristics | 1 | N/A | N/A | N/A | N/A | 0.62/0.59 |
| Boosted Trees | Grouped | Heuristics | 2 | 0.58/0.55 | 0.60/0.57 | 0.61/0.58 | 0.62/0.59 | N/A |
| Boosted Trees | Grouped | Heuristics | 5 | 0.63/0.62 | 0.67/0.66 | 0.71/0.70 | 0.72/0.71 | N/A |
| Boosted Trees | Grouped | Heuristics | 10 | 0.65/0.64 | 0.71/0.70 | 0.74/0.74 | **0.77/0.77** | N/A |
| Boosted Trees | N/A | Bag of Words | 1 | N/A | N/A | N/A | N/A | 0.65/0.61 |
| Boosted Trees | Grouped | Bag of Words | 2 | 0.56/0.52 | 0.60/0.56 | 0.63/0.59 | 0.65/0.61 | N/A |
| Boosted Trees | Grouped | Bag of Words | 5 | 0.63/0.61 | 0.67/0.64 | 0.72/0.70 | 0.76/0.74 | N/A |
| Boosted Trees | Grouped | Bag of Words | 10 | 0.58/0.55 | 0.67/0.65 | 0.76/0.74 | **0.82/0.80** | N/A |
| BERT | N/A | Sequence | 1 | N/A | N/A | N/A | N/A | 0.73/0.73 |
| BERT | Token | Sequence | 2 | 0.86/0.86 | 0.90/0.90 | 0.90/0.90 | **0.95/0.95** | 0.82/0.82 |
| BERT | Token | Sequence | 5 | 0.86/0.86 | 0.89/0.89 | 0.92/0.92 | **0.95/0.95** | 0.80/0.80 |
| BERT | Token | Sequence | 10 | 0.71/0.71 | 0.75/0.76 | 0.81/0.81 | 0.84/0.84 | 0.68/0.68 |
| BERT | N/A | Sequence | 1 | N/A | N/A | N/A | N/A | N/A |
| BERT | Grouped | Sequence | 2 | 0.69/0.67 | 0.69/0.67 | 0.68/0.66 | 0.70/0.68 | N/A |
| BERT | Grouped | Sequence | 5 | 0.71/0.70 | 0.74/0.74 | 0.79/0.78 | 0.83/0.83 | N/A |
| BERT | Grouped | Sequence | 10 | 0.70/0.69 | 0.76/0.76 | 0.82/0.82 | **0.88/0.88** | N/A |

While BERT models outperformed decision tree models overall, the best-performing decision tree model, which was the linguistic-based decision tree with context size 10 utterances and 0% error rate (82%/80% accuracy/F1), outperformed the token-based BERT model trained with context size one utterance and no diarization (73%/73%), and performed on par with the token-based BERT model with two utterances and no diarization. The heuristics-based decision tree model with identical settings also matched the same BERT model in F1 score and accuracy.

### 4.2 Heuristics vs. Lexical Content

Features utilized by the heuristics-based decision tree are shown in Figure 3. First-person pronouns were more associated with patients than medical professionals. Line number was useful for distinguishing between doctors and medical assistants. Linguistic-based boosted decision trees scored higher than heuristics-based boosted decision trees with respect to both accuracy and F1 for each setting of context size and diarization error rate. However, the opposite was the case for the regular decision trees.

**Fig. 3.**
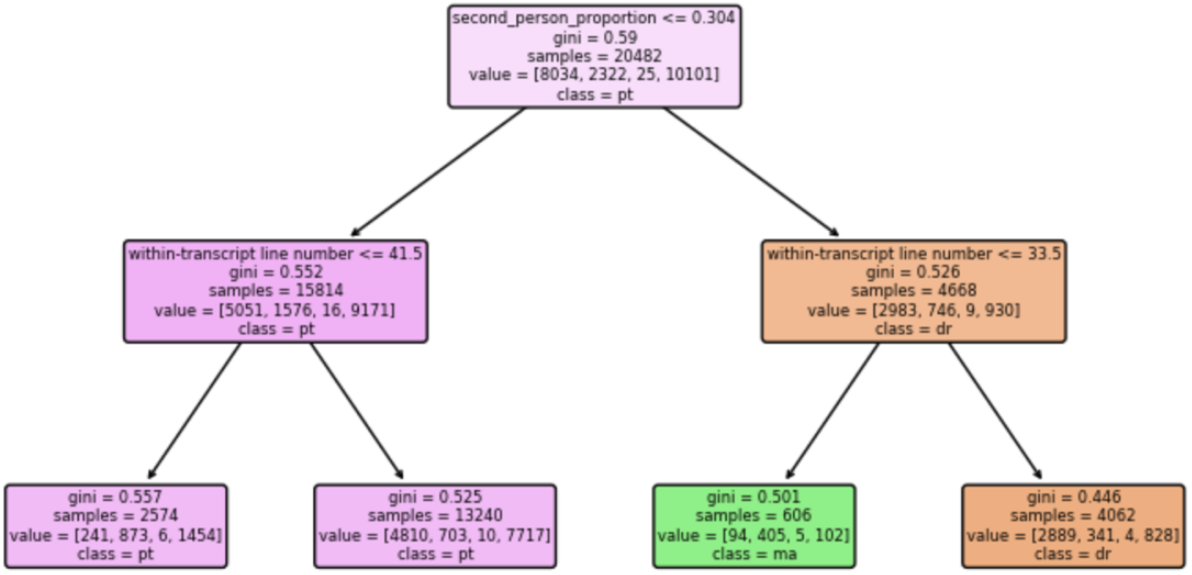
Visualization of Decision Tree trained with 5-utterance context and 100% diarization accuracy. Individual nodes contain the condition (variable and threshold) for moving left (top line), along with the value of the gini purity measure of the applicable records (gini), the number of such records (samples), the ground-truth distribution of the records (value), and the majority class (class).

### 4.3 Diarization-Based Grouping vs. Diarization-Based Tokens

The token-based BERT model outperformed the grouping-based BERT model respect to both accuracy and F1 score at all settings of context size and diarization error rate. The best token-based BERT model achieved both an accuracy and F1 score 95%, whereas the most performant grouping-based BERT model scored 88% on both.

### 4.4 Context Size

Increasing context size beyond a single utterance improved performance of the token-based BERT model. Increasing it to 5 improved performance for some levels of diarization error rate (10%) but decreased it for others (25%, no diarization) and left others unaltered (0%, 50%). However, increasing context size to 10 utterances degraded results. For all other models, increasing context size when diarization error rate was below 50% always increased accuracy and F1 score.

### 4.5 Diarization and Accuracy

Diarization was seen to improve performance for all models when it remained accurate. BERT models which did not use diarization performed uniformly worse than counterparts trained with accurate diarization with respect to accuracy and F1 score. For example, the token-based BERT model with context size 2 scored 82% accuracy, 82% F1 score with no diarization, but achieved a balanced 95%, 90%, 90%, and 86% accuracy and F1 score for settings with 0%, 10%, 25%, and 50% diarization error rate, respectively. Tree-based models also benefited from the combination of context and diarization.

All diarization-based models showed similar patterns of partial reliance on diarization performance, with differences in F1 score between the setting with 0% diarization error rate and 50% diarization error rate ranging from 4% to 18%. There is no clear trend relating the model type and robustness to diarization error.

## 5. Discussion

The purpose of our study was to investigate the potential for LLMs to perform SRI in clinical conversations involving patients, doctors, and medical assistants, and to compare aspects of model design with respect to performance. We find that LLMs can accurately identify who is speaking in clinical conversations using SRI, and furthermore that they benefit from the use of accurate diarization despite remaining robust to errors. This contributes to the growing literature on SRI by demonstrating its possibility in clinical settings and providing helpful comparisons of design considerations. Despite being the first of their kind for clinical SRI in clinical conversations, our models perform on par with recent text transformer-based SRI models trained in other domains such as Air Traffic Control [11].

Comparison of boosted decision tree models and LLMs reveal that model class makes a large difference in downstream performance, with LLM models outperforming boosted decision tree models for SRI in clinical conversations. Comparison of decision tree models and boosted decision trees reveals the same finding. This is likely a result of the fact that boosted decision trees can approximate a wider range of functions than decision trees, and LLMs a yet wider range, allowing them to model the intricate semantic dependencies between speaker role and conversational utterances with increasing intricacy. LLM superiority likely also results from the fact that LLM embeddings encode richer semantic information than bag-of-words features, including positional information and synonymy relations.

Comparison of the boosted decision tree trained using second-person pronoun proportions versus using bag-of-words reveals that pronoun-usage heuristics underperform lexical approaches for SRI in our clinical conversation dataset. This is because the latter permits learning a more general set of role-specific keywords, including but not limited to tendencies in pronoun usage. Nevertheless, pronoun usage trends are important for performance in lexical approaches: presence of the word “you” serves as the primary split in all our learned decision trees. The fact that decision trees performed better with heuristics is not evidence that lexical approaches underperform heuristics due to the tendency for decision trees to overfit in the presence of complex data types such as text [20].

LLM models which employed diarization outperformed LLM models which did not, as long as diarization identifiers remained accurate, indicating that diarization identifiers can be useful as input signals to SRI pipelines, as long as accuracy is maintained. Decision trees which were able to attend to context in the presence of diarization identifiers outperformed single-utterance decision trees as well, demonstrating that use of diarization-based grouping methods can allow decision trees to benefit from larger context sizes effectively.

Amongst LLM models which attended to diarization, the grouping-based models underperformed relative to the token-based models. This indicates that models benefit from being able to flexibly attend to diarization signals rather than being constrained by them to analyze utterance groups which may or may not be malformed due to diarization errors.

For all grouping-based models, increasing context size had a positive impact on performance. However, for the LLM models trained without diarization-based grouping, increasing context past a certain point had the tendency to degrade performance. Interestingly, this trend persisted in the absence of diarization. This suggests that excessive context has the tendency to degrade results when adequate filtering steps are not taken to minimize input sizes. Despite potential inaccuracies, diarization-based grouping can act as a filtering mechanism to reduce input token sizes.

All models which used diarization benefited from increased diarization accuracy. Interestingly, the token-based LLM did not have a more robust performance against diarization inaccuracy compared to grouping-based methods, indicating that diarization errors may propagate through LLMs regardless of whether grouping is used or not.

## 6. Limitations

While the current study has revealing findings concerning design and implementation of SRI systems in clinical settings, there exist limitations which will need to be addressed in future works. First, our analysis was limited to a single dataset of clinical conversations, leaving questions about generalizability across specialty and institution. Second, our experiments only simulated one aspect of diarization error; namely, when diarization identifiers are inconsistent. However, future experiments should also simulate errors in segmentation due to voice activity detection, which is another component of diarization. Lastly, our experiments only use one candidate LLM, which can potentially limit generalizability to other models. However, given the number of open-source LLMs and the fact that many of them are comparable to and more performant than BERT in a wide array of NLP tasks, we hypothesize that our results are broadly applicable to performant LLMs and may even act as a lower bound on possible performance.

## 7. Conclusion

We demonstrate that semantic cues alone can identify speaker roles in clinical conversations. These are more powerful than specific communication heuristics. While diarization is useful for various model types depending on how it is incorporated, our LLM-based approach is robust to diarization errors and even remains performant in the absence of diarization. Further, non-LLM based approaches which use simple techniques such as bag-of-words also perform well in the context of clinical SRI and remain somewhat robust to diarization errors. Lastly, LLM approaches which attend to a target utterance in the context of previous utterances enriched with diarization labels are shown to be superior to utterances which analyze disjoint groups of diarized utterances separately, whether diarization is accurate. These findings support the viability of SRI pipelines for clinical interaction analysis using transcribed conversations and have implications for the implementation of conversational analysis in clinical systems. Applications of this technology allow speaker role-based segmentation of visits at scale, providing a possible foundation for downstream modeling, such as speaker-based sentiment analysis and measurement of communication quality for clinical professionals.

## Data Availability

Due to sensitivity of data, data is not publicly available.

## References

1. Fawole, Oluwakemi A. et al. “A systematic review of communication quality improvement interventions for patients with advanced and serious illness.” Journal of general internal medicine vol. 28,4 (2013): 570–7. doi:10.1007/s11606-012-2204-4

2. Crampton, Noah H. et al. “Computers in the clinical encounter: a scoping review and thematic analysis.” Journal of the American Medical Informatics Association : JAMIA vol. 23,3 (2016): 654–65. doi:10.1093/jamia/ocv178

3. Beck, Rainer S. et al. “Physician-patient communication in the primary care office: a systematic review.” The Journal of the American Board of Family Practice vol. 15,1 (2002): 25–38.

4. Kavic, Michael S. “Competency and the six core competencies.” JSLS : Journal of the Society of Laparoendoscopic Surgeons vol. 6,2 (2002): 95–7.

5. Nasca, Thomas J et al. “The next GME accreditation system--rationale and benefits.” The New England journal of medicine vol. 366,11 (2012): 1051–6. doi:10.1056/NEJMsr1200117

6. Epstein, Ronald M., and Richard L. Street Jr. “The values and value of patient-centered care.” Annals of family medicine vol. 9,2 (2011): 100–3. doi:10.1370/afm.1239

7. Brockway, Christine. “Evaluating Effective Communication in Clinical and Simulation.” Clinical Simulation in Nursing, vol. 98, Jan. 2025, article no. 101669. 10.1016/j.ecns.2024.101669.

8. Park, Tae Jin, et al. “A Review of Speaker Diarization: Recent Advances with Deep Learning.” Computer Speech & Language, vol. 72, Mar. 2022, article no. 101317, 10.1016/j.csl.2021.101317.

9. Nghiem, Minh-Quoc, et al. “Speaker Role Identification in Call Centre Dialogues: Leveraging Opening Sentences and Large Language Models.” Proceedings of the 24th Annual Meeting of the Special Interest Group on Discourse and Dialogue, edited by Svetlana Stoyanchev et al., Association for Computational Linguistics, 2023, pp. 388–392. 10.18653/v1/2023.sigdial-1.35.

10. O’Sullivan, James et al. “Automatic speaker diarization for natural conversation analysis in autism clinical trials.” Scientific reports vol. 13,1 10270. 24 Jun. 2023, doi:10.1038/s41598-023-36701-4

11. Guo, Dongyue, et al. “A Comparative Study of Speaker Role Identification in Air Traffic Communication Using Deep Learning Approaches.” ACM Transactions on Asian and Low-Resource Language Information Processing, vol. 22, no. 4, Apr. 2023, article 102, 10.1145/3572792.

12. Zolnoori, Maryam et al. “Is the patient speaking or the nurse? Automatic speaker type identification in patient-nurse audio recordings.” Journal of the American Medical Informatics Association : JAMIA vol. 30,10 (2023): 1673–1683. doi:10.1093/jamia/ocad139

13. Cheema, Gullal S., et al. “Identification of Speaker Roles and Situation Types in News Videos.” Proceedings of the 2024 International Conference on Multimedia Retrieval, Association for Computing Machinery, 2024, pp. 506–514. 10.1145/3652583.3658101.

14. Barzilay, Regina, et al. “The Rules Behind Roles: Identifying Speaker Role in Radio Broadcasts.” Proceedings of the Seventeenth National Conference on Artificial Intelligence and Twelfth Conference on Innovative Applications of Artificial Intelligence, AAAI Press, 2000, pp. 679–684.

15. Hutchinson, Brian, et al. “Unsupervised Broadcast Conversation Speaker Role Labeling.” 2010 IEEE International Conference on Acoustics, Speech and Signal Processing, Dallas, TX, USA, 2010, pp. 5322–5325. IEEE. doi:10.1109/ICASSP.2010.5494958.

16. Sapru, Ashtosh, and Fabio Valente. “Automatic Speaker Role Labeling in AMI Meetings: Recognition of Formal and Social Roles.” 2012 IEEE International Conference on Acoustics, Speech and Signal Processing (ICASSP), Kyoto, Japan, 2012, pp. 5057–5060. IEEE. doi:10.1109/ICASSP.2012.6289057

17. Robins L, Brock D, Mauksch L. Effects of Establishing Focus in the Medical Interview. Final Progress Report. AHRQ Grant Number R01HS013172 (PI: Robins, L). Retrieved from: https://www.ahrq.gov/sites/default/files/2024-07/robins-report.pdf

18. Devlin, Jacob, et al. “BERT: Pre-training of Deep Bidirectional Transformers for Language Understanding.” Proceedings of the 2019 Conference of the North American Chapter of the Association for Computational Linguistics: Human Language Technologies, vol. 1, 2019, pp. 4171–4186. Association for Computational Linguistics.

19. Vaswani, Ashish, et al. “Attention is All You Need.” Proceedings of the 31st International Conference on Neural Information Processing Systems, Long Beach, California, 2017, Curran Associates Inc., pp. 6000–6010.

20. Halabaku, Erblin, and Eliot Bytyçi. “Overfitting in Machine Learning: A Comparative Analysis of Decision Trees and Random Forests.” Intelligent Automation & Soft Computing, vol. 39, no. 6, 2024, pp. 987–1006. 10.32604/iasc.2024.059429.

